# Beyond the Checklist: A large-scale analysis of under-recognised weight loss behaviours in individuals with eating disorders

**DOI:** 10.1101/2025.02.17.25322397

**Authors:** Saakshi Kakar, Una Foye, Helena L. Davies, Elisavet Palaiologou, Chelsea M. Malouf, Laura Meldrum, Iona Smith, Gursharan Kalsi, Karina L. Allen, Gerome Breen, Moritz Herle, Christopher Hübel

## Abstract

**Objective:** This study aimed to identify and categorise under-recognised weight loss behaviours in individuals with eating disorders, addressing diagnostic gaps.

**Method:** We text mined free-text responses from 1,675 participants with anorexia nervosa, bulimia nervosa, or binge-eating disorder in the Genetic Links to Anxiety and Depression (GLAD) Study and the United Kingdom Eating Disorders Genetics Initiative (EDGI UK). In secondary analyses, we investigated differences by eating disorder and gender.

**Results:** Frequently endorsed behaviours included structured diets (341 endorsements) and calorie counting (321 endorsements) but also less commonly considered behaviours like compression garments (113 endorsements) and self-harm (63 endorsements). We identified four overarching themes: restriction-based approaches, medical intervention, body manipulation, and food avoidance. The most frequently reported weight loss behaviours and resultant themes did not differ among eating disorders or genders, closely resembling those in the broader sample.

**Discussion:** Our findings identify a crucial gap in current diagnostic criteria, which may hamper recognition and lead to underdiagnosis of eating disorders. By incorporating our insights, clinicians could capture a broader spectrum of behaviours, thus improving diagnostic accuracy. However, our sample homogeneity implicates the need for more diverse samples. Our study contributes essential insights for enhancing diagnostic criteria.

**Public Significance Statement:** Our study revealed under-recognised weight loss behaviours in people with eating disorders that current diagnostic tools miss, which may lead to underdiagnoses. By identifying these behaviours and taking a broader diagnostic approach, our research can help clinicians better understand eating disorders by improving diagnostic accuracy and opening up new avenues for personalised care.

## Introduction

Weight loss behaviours are used to try to control body weight or shape (Loth et al., 2014) and constitute core symptoms of many eating disorders. The Diagnostic and Statistical Manual of Mental Disorders, fifth edition (DSM-5) lists six weight loss behaviours for bulimia nervosa: non-purging behaviours: fasting, excessive exercise, diet pill use; and, purging behaviours: self-induced vomiting, laxative use, and diuretic use (Abebe et al., 2012; American Psychological Association, 2013). These behaviours may be present in anorexia nervosa and other specified feeding or eating disorders (OSFED). While purging behaviours are often undertaken to reduce caloric intake, they are largely ineffective at this, particularly in the case of laxatives and diuretics, whose impact on calorie absorption is minimal (Juarascio et al., 2018). Weight loss behaviours are associated with physical complications, such as cardiomyopathy and rectal prolapse, and psychiatric disorders, including major depressive disorder, generalised anxiety disorder, and suicidality (Campbell & Peebles, 2014; Spindler & Milos, 2007; Westmoreland et al., 2016).

Eating disorder assessment tools vary in the scope of evaluated behaviours. The ED100K.V3 (Bulik et al., 2021) includes DSM-5 listed behaviours for bulimia nervosa. Conversely, the Eating Disorder Examination Questionnaire (EDE-Q 6.0; Fairburn & Beglin, 1994) and the Eating Pathology Symptoms Inventory (EPSI; Forbush et al., 2013), two widely recognised assessment tools used in both research and clinic, assess a broader range. The EDE-Q captures dietary restriction tied to weight and shape concerns, while the EPSI evaluates excluding “unhealthy” and high-calorie foods from one’s diet, muscle-building supplements, protein supplements, and diet teas. Current assessment tools are limited by their range of assessed weight loss behaviours and their underlying motivations, such as the intent to lose weight.

Weight loss behaviours are prevalent in the general population, even in individuals as young as 12 years old (Forman-Hoffman, 2004; Gonsalves et al., 2014; Neumark-Sztainer et al., 2006; Solmi et al., 2021). In the US, 34% of middle school students reported using weight loss methods (Yeatts et al., 2016). Adolescent girls who experienced and were bothered by weight-based teasing reported significantly more weight loss behaviours both at baseline and during a 10-year follow-up compared to those who were not teased (Simone et al., 2019).

### Non-purging weight loss behaviours

Fasting is characterised as voluntarily restricting food and drink intake for longer than an overnight fasting period. It can be categorised based on timing: alternate-day fasting, time-restricted feeding, and whole-day fasts (Visioli et al., 2022). Among 2,856 Australian adults in a community sample, 1.9% of men and 2.4% of women reported fasting at least thrice weekly to lose weight (Bentley et al., 2014). In stark contrast, ∼70% of participants with anorexia nervosa from Australia and New Zealand reported engaging in fasting to control their weight (Kirk et al., 2017).

Excessive exercise plays a crucial role in the aetiology of eating disorders (Fietz et al., 2014). Definitions of excessive exercise vary across studies. The DSM-5 defines it as a behaviour disrupting essential activities, occurring at unsuitable times or settings, or persisting despite injuries or medical complications (American Psychological Association, 2013; Mond & Gorrell, 2021). In a U.S.-American community sample, 54% of men and 52% of women reported engaging in excessive exercise (Kruger et al., 2004), while its estimated prevalence in the general adult population in Hungary is ∼0.5% (Mónok et al., 2012). Among individuals with anorexia nervosa, >80% reported exercising to control their body weight, with an average onset at 16 years (Kirk et al., 2017). In a systematic review, the prevalence of excessive exercise among adolescents with eating disorders ranged from 16.7 to 85.3% (Fietz et al., 2014).

Finally, diet medications like liraglutide, orlistat, and the increasingly popular semaglutide are used to control body weight or shape (Suran, 2023). Moreover, amongst American college students, 2.9% reported lifetime use of nonprescription diet pills for weight control, with Hispanic/Latin and Biracial/Multiracial students more likely to take diet pills than white students (Van Dyne et al., 2023). Diet pill use is common among individuals with eating disorders, with the highest prevalence reported in those with the purging subtype of anorexia nervosa (41%) and bulimia nervosa (44%; Reba-Harrelson et al., 2008), while women with the anorexia nervosa restricting subtype (8%) use them significantly less frequently than those with other eating disorders. Of nearly 4,000 participants with anorexia nervosa, ∼17% frequently used diet pills to control their weight (Kirk et al., 2017).

### Purging weight loss behaviours

While self-induced vomiting is perhaps the most recognised purging behaviour, a systematic review indicates that the point prevalence of vomiting varies widely, ranging from less than 1 to 11% of the adult general population, depending on the country (Ortega-Luyando et al., 2015). In comparison, two studies found that 31-56% of patients with eating disorders in a treatment facility used self-induced vomiting as a weight loss behaviour (Dalle Grave et al., 2009; Johannson et al., 2022). In Australasia, 40% of participants with anorexia nervosa frequently engaged in self-induced vomiting (Kirk et al., 2017). Additionally, amongst 159 participants with eating disorders with binge eating, white participants significantly more likely engaged in self-induced vomiting (29%) than Black participants (3%; Lin et al., 2022).

Laxatives are widely available over-the-counter medications which stimulate bowel movements (Werth & Christopher, 2021). Lifetime use of laxatives for weight control among U.S.-American adults is estimated at 5% (Levinson et al., 2020). The prevalence of laxative use in those with eating disorders is much higher with estimates between 10 and 60% (Roerig et al., 2010; Steffen et al., 2007).

Lastly, diuretics, known as “water pills”, are prescription medications aiding in the elimination of excess water by increasing urine production (Ellison, 2019). Out of a nationally representative sample of 9,419 U.S. participants, 1.4% of men and 2% of women used diuretics to try to lose weight (Kruger et al., 2004). The use of diuretics for weight control is more frequent in those with eating disorders, particularly bulimia nervosa with 31% compared with 5.5% in anorexia nervosa (Huas et al., 2011; Roerig et al., 2003).

### Weight loss behaviours not captured by the DSM-5

Few studies have examined weight loss behaviours in individuals with eating disorders beyond those included in the DSM-5. For example, individuals with self-reported eating disorders endorse vaping for appetite and weight control more often than those without eating disorders (Morean & Insalata, 2017). Chewing and spitting was reported by a quarter of 359 individuals with an eating disorder (Song, Lee & Jung, 2015). Nevertheless, these studies may scratch the surface of the complexity of weight loss behaviours, suggesting a misrepresentation of weight loss behaviours in diagnostic criteria. Additionally, young people turn to social media to understand their own behaviours, seek help, and find a sense of inclusion and validation (Lavis & Winter, 2020).

Weight loss behaviours increase health risks in anyone engaging in them. Research and clinical practice are limited by diagnostic criteria. Qualitative research enables in-depth identification of unknown behaviours, uncovering a broader range and providing deeper insights into individuals’ experiences and unmet needs. We aimed to identify and categorise weight loss behaviours in individuals with eating disorders, using qualitative analysis of free-text responses, and hence contribute to developing more inclusive and effective assessment materials.

## Methods

### Study design and sample

Data were collected as part of the Genetic Links to Anxiety and Depression (GLAD; gladstudy.org<u>.uk</u>) Study and The United Kingdom Eating Disorders Genetics Initiative (EDGI UK; edgiuk.org). Both are ongoing online studies collecting genetic and phenotypic data from UK-based individuals over 16 years old. The studies form part of the National Institute for Health and Care Research (NIHR) Mental Health BioResource; a data resource comprising recontactable participants.

The GLAD Study investigates the genetic and environmental bases of anxiety and depressive disorders (Davies et al., 2019). Participants live in the United Kingdom and either have a lifetime experience of anxiety and/or depressive disorders (cases) or have never experienced any mental health conditions (controls). Present analyses comprise responses from cases to the optional ED100K questionnaire, collected on or before 16 September 2022 (Thornton et al., 2018).

EDGI UK launched in February 2020 to recruit 10,000 participants who have ever experienced an eating disorder (Monssen et al., 2023). Participants are residents of England with a lifetime experience of eating disorders. Present analyses consist of all sign-up questionnaires completed on or before 16 September 2022.

The London - Fulham Research Ethics Committee approved the GLAD Study on 21st August 2018 (REC reference: 18/LO/1218) and EDGI UK on 29th July 2019 (REC reference: 19/LO/1254).

### Measures

Demographic characteristics (age, sex assigned at birth, gender, ethnicity, highest level of education, sexuality) were collected through the GLAD Study and EDGI UK sign-up questionnaires.

Lifetime eating disorder diagnoses were self-reported in the GLAD Study and EDGI UK via the question, *“Have you been diagnosed with one or more of the following mental health problems by a professional, even if you don’t have it currently?”* with eating disorders as response options as listed in the DSM-5 (American Psychological Association, 2013; Davies et al., 2020).

Eating disorder diagnoses were additionally determined using diagnostic algorithms, based on responses to the ED100K.V3, using DSM-5 criteria (American Psychological Association, 2013; Bulik et al., 2021; github.com/tnggroup/EDGI_protocol). The ED100K.V3 is a self-report questionnaire that assesses lifetime eating disorder symptoms (Bulik et al., 2021), based on the Structured Clinical Interview for DSM-5, Eating Disorders. The ED100K-V1 is clinically validated for criteria B and C for anorexia nervosa and for binge eating (Thornton et al., 2018). Whilst an optional questionnaire in the GLAD Study, the ED100K.V3 is the basis of the sign-up questionnaire in EDGI UK. A hierarchical categorisation (anorexia nervosa restricting > anorexia nervosa binge-eating/purging > anorexia nervosa unknown subtype > bulimia nervosa > binge-eating disorder) was applied to ensure no overlapping cases among the algorithm-defined eating disorders. This hierarchy is common practice in epidemiological studies (Schaumberg et al., 2019; Udo & Grilo, 2018). For inclusion, participants must have answered the ED100K.V3 (Bulik et al., 2021) and have either a self-reported or an algorithm-derived eating disorder: anorexia nervosa, bulimia nervosa, or binge-eating disorder.

In the ED100K.V3, the free-text question: *“Have you ever used any other methods not listed in the previous question to control your body shape or weight”* assessed weight loss behaviours and compensatory behaviours, used to offset binge eating, were assessed via: *“Please state any other methods used to compensate for episodes of binge eating or overeating.”* (Bulik et al., 2021).

Some participants provided details about their weight loss behaviours, which have been paraphrased to protect participant confidentiality and are highlighted in italics in the results section.

### Data analyses

Prior to analyses, we merged data from the GLAD Study and EDGI UK ED100K.V3, cleaned it, and reported descriptives (**Table 1**). Analyses were conducted using the R package *tidytext* (Silge & Robinson, 2016). We text mined (i.e., extracted patterns from text) to analyse free-text responses (**Figure 1**; Jo, 2019). We used a thematic analysis approach (Braun & Clarke, 2006). We made interpretation and coding decisions rather than relying on automated processes. This qualitative nature of our work ensures that the identified themes reflect the context and meaning expressed by the participants.

**Figure 1.**
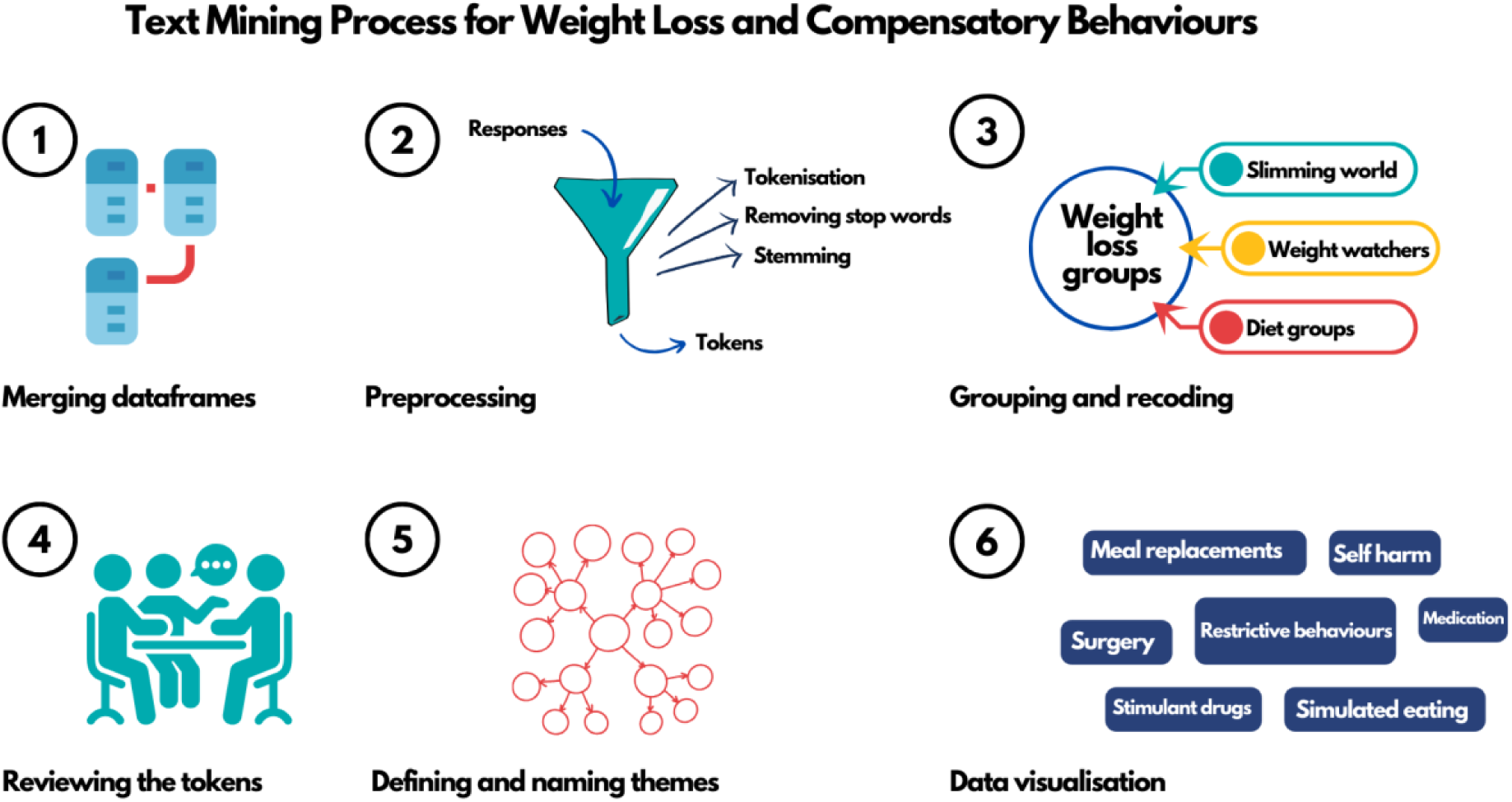
Analysis flowchart of text mining on free-text responses to weight loss and compensatory behaviours questions from participants of the GLAD Study and EDGI UK.

**Table 1.** Demographic characteristics of the GLAD and EDGI UK sample that answered the ED100K.V3 (*n* = 1675). Eating disorder diagnoses are either algorithm-derived or self-reported. The unknown subtype of anorexia nervosa results from the self-report questionnaire not differentiating between subtypes in the GLAD study. Due to low numbers, we created a merged category “Black, Black British, or Arab” to protect privacy.

| Characteristic | n | % |
| --- | --- | --- |
| Sex |  |  |
| Female | 1588 | 94.8% |
| Male | 87 | 5.2% |
| Gender |  |  |
| Woman | 1510 | 90.2% |
| Man | 99 | 5.9% |
| Non-binary | 42 | 2.5% |
| Self-defined | 13 | 0.8% |
| Don’t know/Prefer not to say | 11 | 0.7% |
| Eating disorder diagnosis |  |  |
| Anorexia nervosa |  |  |
| Restricting | 231 | 13.8% |
| Binge-eating/purging | 452 | 27.0% |
| Unknown subtype | 8 | 0.5% |
| Bulimia nervosa | 903 | 53.9% |
| Binge-eating disorder | 81 | 4.8% |
| Ethnicity |  |  |
| White | 1568 | 93.6% |
| Mixed | 61 | 3.6% |
| Asian or Asian British | 17 | 1.0% |
| Black, Black British, or Arab | 8 | 0.5% |
| Don't know/Other | 21 | 1.1% |
| Highest level of education |  |  |
| University | 825 | 49.3% |
| A-levels/AS-levels or equivalent | 487 | 29.1% |
| NVQ/HND/HNC or equivalent | 103 | 6.2% |
| GCSEs/CSEs/O-levels or equivalent | 200 | 11.9% |
| None of the above | 42 | 2.5% |
| Seen but not answered | 18 | 1.1% |
| Sexuality |  |  |
| Heterosexual | 1024 | 61.1% |
| Homosexual | 96 | 5.7% |
| Bisexual | 389 | 23.2% |
| Asexual | 40 | 2.4% |
| Self-defined | 48 | 2.9% |
| Other | 46 | 2.8% |
| Prefer not to answer | 32 | 1.9% |

**1. Merging data frames:** We merged data from the GLAD Study and EDGI UK, creating separate data frames for responses discussing weight loss behaviours and compensatory behaviours (i.e., following binge eating).

**2. Preprocessing**: We performed preprocessing operations to prepare the data frames for analysis.

**2.1. Tokenisation**: Splitting the text into meaningful units (in this case, sentences).

**2.2. Stop word removal**: Eliminating words like “a” and “the” that do not have semantic value for the analysis.

**2.3 Stemming**: Reducing words to their root form.

**3. Categorising and recoding**: We categorised and recoded sentence tokens referencing the same or similar weight loss and compensatory behaviours into codes. For instance, responses like “slimming world” and “weight watchers” were combined and recoded as “weight loss groups”.

**3.1. Handling multiple behaviours in a token**: When a participant endorsed more than one behaviour in a token, the responses were recoded with a full stop between meaningful chunks to allow separate tokenisation and individual analysis. For example, “Smoking and self-harm” was recoded to “Smoking. Self-harm.”, often resulting in more than one token per participant.

**3.2. Combining multi-sentence behaviours**: When participants described one behaviour across multiple sentence tokens, the responses were recoded into a single token to avoid double-counting. For example, “Chew gum instead of food. Excessively” was recoded to “Chew gum instead of food”.

**3.3. Calculating endorsement of behaviour tokens:** To determine the percentage of endorsement for each weight loss and compensatory behaviour, we calculated the proportion of tokens endorsing a specific behaviour relative to the total number of response tokens per question.

**4. Reviewing the tokens:** The initial coding was performed by the first author, followed by a second round by EP and HLD using Excel. Any discrepancies were resolved through team discussions.

**5. Defining and naming themes:** We organised the tokens into potential themes, and categorised related codes together to form cohesive and meaningful categories that accurately represent the weight loss and compensatory behaviours reported by participants. While some behaviours may fit into multiple themes, we categorised them based on their most prominent characteristics—such as their primary function or intended outcome— to maintain clarity and analytical focus.

**6. Data visualisation:** We then created graphics to visually represent the most endorsed behaviour themes.

In secondary analyses, the text mining process outlined above was repeated to examine differences by eating disorder and gender.

1. **Data Segmentation**: The dataset was filtered to isolate responses from participants with anorexia nervosa, bulimia nervosa, and binge-eating disorder and separately by gender. The subsets were then subjected to the same preprocessing steps as the overall dataset.
2. **Behavioural Categorisation**: The same categorisation and recoding strategies were applied to categorise behaviours within these subgroups, ensuring consistency with the initial analysis.
3. **Interpretation of Results**: The results of this secondary analysis were compared to the overall findings, providing insights into how individuals with different eating disorder diagnoses and men may exhibit distinct behaviours within the context of the study.

## Results

The total sample consisted of 1,675 participants, 94.8% female, from the GLAD Study and EDGI UK (**Table 1**) aged on average 31.7 years (SD = 13.1; range = 16–77). In the sample, 27% had a diagnosis of anorexia nervosa binge-eating/purging and 13.8% had a restricting subtype. Additionally, 53.9% of participants had a bulimia nervosa diagnosis and 4.8% binge-eating disorder.

After excluding missing data (i.e., NA and “Seen but not answered”; 13,386 participants did not respond to the weight loss behaviour question and 16,563 participants did not respond to the compensatory behaviour question) and preprocessing, 3,492 tokens regarding weight loss behaviours and 815 tokens regarding compensatory behaviours were available (**Table 2**). The high number of nonresponses to the free-text questions likely reflects the greater cognitive and time demands than Likert scale questions.

**Table 2.** Frequency of endorsed weight loss behaviours (*n* = 3,492 tokens) and compensatory behaviours (*n* = 815 tokens), as well as weight loss behaviours reported by men (*n* = 99 tokens), based on free-text responses from the ED100K questionnaire in the GLAD Study and EDGI UK.

| **Behaviour tokens** | **Weight loss behaviour endorsement ($n = 3,492$)** | **Compensatory behaviour endorsement ($n = 815$ tokens)** | **Men’s endorsement ($n = 99$ tokens)** |
| --- | --- | --- | --- |
| Structured/popular weight loss diets, e.g., Atkins® diet, Cambridge diet | 619 (17.7%) | 421 (51.7%) | 16 (16.1%) |
| Calorie counting | 422 (12.1%) | 281 (34.5%) | 7 (7.1%) |
| Restrictive eating, e.g., restrictive food rules | 389 (11.1%) | 279 (34.2%) | 10 (10.1%) |
| Chewing & spitting | 214 (6.1%) | 155 (19.0%) | <5 |
| Illicit drugs, e.g., cocaine, amphetamines | 213 (6.1%) | 154 (19.0%) | 12 (12.1%) |
| Fluid loading | 207 (5.9%) | 94 (11.5%) | <5 |
| Weight loss groups, e.g., slimming world | 160 (4.6%) | 136 (16.7%) | <5 |
| Meal replacements, e.g., meal replacement milkshakes | 155 (4.4%) | 117 (14.4%) | <5 |
| Compression garments, e.g., corsets | 147 (4.2%) | 73 (9.0%) | <5 |
| Bariatric surgery, e.g., gastric bypass, gastric bands | 146 (4.2%) | 100 (12.3%) | 6 (6.1%) |
| Appetite suppressants, e.g., caffeine, smoking | 122 (3.5%) | 74 (9.1%) | 5 |
| Self-harm | 88 (2.5%) | 49 (6.0%) | <5 |
| Prescription medication, e.g., metformin | 81 (2.3%) | 59 (7.2%) | 6 (6.1%) |
| Reducing body temperature, e.g., ice baths | 68 (1.9%) | 23 (2.8%) | <5 |
| Inducing sweating | 47 (1.3%) | 33 (4.0%) | 0 |
| Consuming foods incompatible with intolerances | 47 (1.4%) | 21 (2.6%) | <5 |
| Cognitive methods, e.g., engaging with pro-anorexia/pro-bulimia websites, body checking, hypnotherapy, mindfulness, negative imagery | 45 (1.3%) | 9 (1.1%) | 0 |
| Hiding/disposing of food | 37 (1.1%) | 23 (2.8%) | 0 |
| Restricting fluid intake | 33 (0.9%) | 21 (2.6%) | 0 |
| Micro-exercise, e.g., leg tapping, pacing, fidgeting | 31 (0.9%) | 0 | 0 |
| Sleeping to avoid eating | 31 (0.9%) | 15 (1.8%) | 0 |
| Eating non-food items, e.g., tissue, wallpaper paste, cotton balls | 29 (0.8%) | 15 (1.8%) | <5 |
| Excessive alcohol consumption | 26 (0.7%) | 12 (1.5%) | <5 |
| Chewing gum | 22 (0.6%) | 12 (1.5%) | 0 |
| Colon cleansing, e.g., colonic irrigation, enemas | 18 (0.5%) | 14 (1.7%) | <5 |
| Electrical muscle stimulation | 17 (0.5%) | 10 (1.2%) | <5 |
| Weight loss injections, e.g., saxenda | 13 (0.4%) | 16 (2.0%) | 0 |
| Manipulating a nasogastric tube | 11 (0.3%) | 16 (2.0%) | 0 |
| Sleep deprivation | 6 (0.2%) | 0 | 0 |
| Overeating to induce vomiting | 5 (0.1%) | 0 | 0 |
| Energy drinks | 5 (0.1%) | 0 | 0 |
| Steroids | 5 (0.1%) | 0 | 0 |

The weight loss and compensatory behaviour tokens were highly similar. Therefore, we categorised tokens into four themes providing a more structured understanding of the weight control methods: restriction-based approaches, medical interventions, body manipulation, and food avoidance (**Figure 2**). These themes highlight the range of strategies used to control weight, from more common dietary methods to less common, more extreme practices not typically captured by standard assessments.

**Figure 2.**
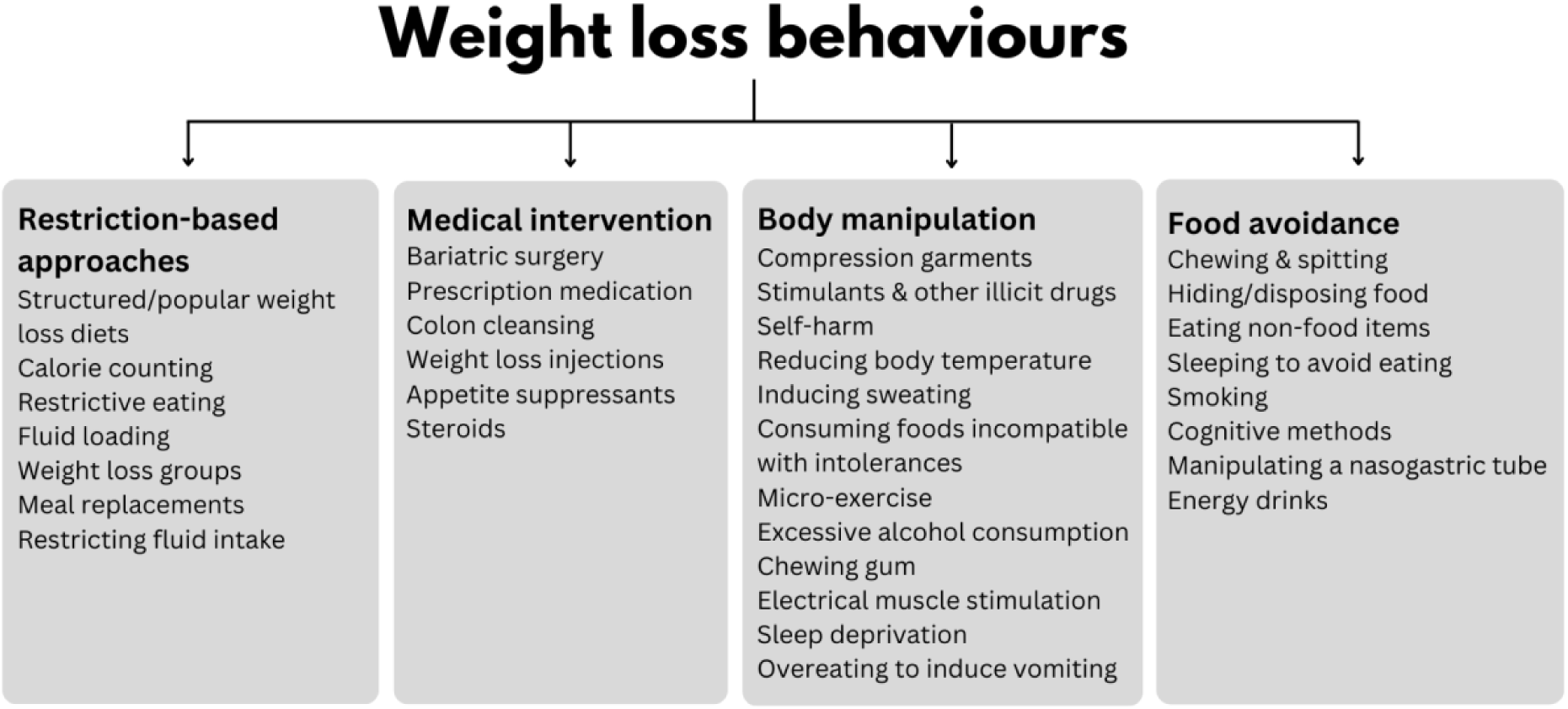
The identified themes from respondents’ weight loss behaviours.

Restriction-based approaches encompass behaviours focused on controlling food and fluid intake including diet-based strategies, such as structured/popular weight loss diets like the Atkins^®^ Diet, and participation in weight loss groups like WeightWatchers^®^. Additionally, fluid-related behaviours, such as consuming excessive amounts of water or carbonated drinks to create a *sense of fullness*, or conversely, restricting fluid intake to avoid feeling bloated and *with the fear it would make them fat*, were noted. Many participants reported engaging in a combination of behaviours, resulting in several tokens.

Medical interventions include invasive or pharmaceutical methods including bariatric surgery, such as gastric bypass and gastric bands, which restricts the stomach’s capacity to hold food. Additionally, participants endorsed using medications like metformin (used to treat type 2 diabetes), methylphenidate (used to treat attention-deficit/hyperactivity disorder), and levothyroxine (used to treat hypothyroidism) for weight loss rather than their prescribed intent. Participants reported using various substances, including caffeine from coffee or pills, illicit drugs, and smoking to suppress appetite.

Body manipulation captures physically manipulative methods. Participants described lowering their body temperature through baths, intentionally keeping cold, or inducing sweating by wrapping themselves in cling film or spending extended periods in saunas to *use up calories*. Some individuals engaged in self-harm, such as cutting or burning fat off their bodies, as a means to directly reduce body weight. Some deliberately consumed foods incompatible with their intolerances, such as eating gluten despite being coeliac, to provoke physical discomfort that discouraged eating, served as a form of *punishment* to suppress food intake, or was believed to expend calories. Some participants also engaged in sleep deprivation, viewing it as a strategy to burn calories, while others reported overeating deliberately to induce vomiting as a means of purging. Other methods included engaging in “micro-exercises,” performing small, repetitive movements throughout the day to burn calories without drawing attention. Excessive alcohol consumption was also mentioned as a tactic to reduce food intake and *make them feel sick*. Finally, respondents wore compression garments, like corsets or waist trainers, to physically restrict the stomach, create a slimmer appearance, and make eating *uncomfortable*.

Food avoidance encompasses behaviours to prevent or reduce food consumption, such as chewing and spitting and, in more extreme cases, consuming non-food items to increase stomach distension without ingesting calories. Chewing gum was frequently used to create a sensation of fullness, suppress hunger, or provide a distraction from cravings, potentially leveraging the cephalic phase response to simulate the experience of eating without caloric intake.

In the secondary analysis, we analysed responses separately for anorexia nervosa (across subtypes; *n* = 691), bulimia nervosa (*n* = 903), and binge-eating disorder (*n* = 81; **Figure 3**). Amongst participants with anorexia nervosa, the three most frequently reported behaviours were calorie counting (13.3%), chewing and spitting (12.7%) and restrictive eating (12.3%). For participants with bulimia nervosa, the top three behaviours were structured/popular weight loss diets (18.1%), calorie counting (9.5%), and meal replacements (8.5%). Amongst participants with binge-eating disorder, structured/popular weight loss diets (16.1%), restrictive eating (14.8%), and bariatric surgery (13.6%) were the most frequently reported.

**Figure 3.**
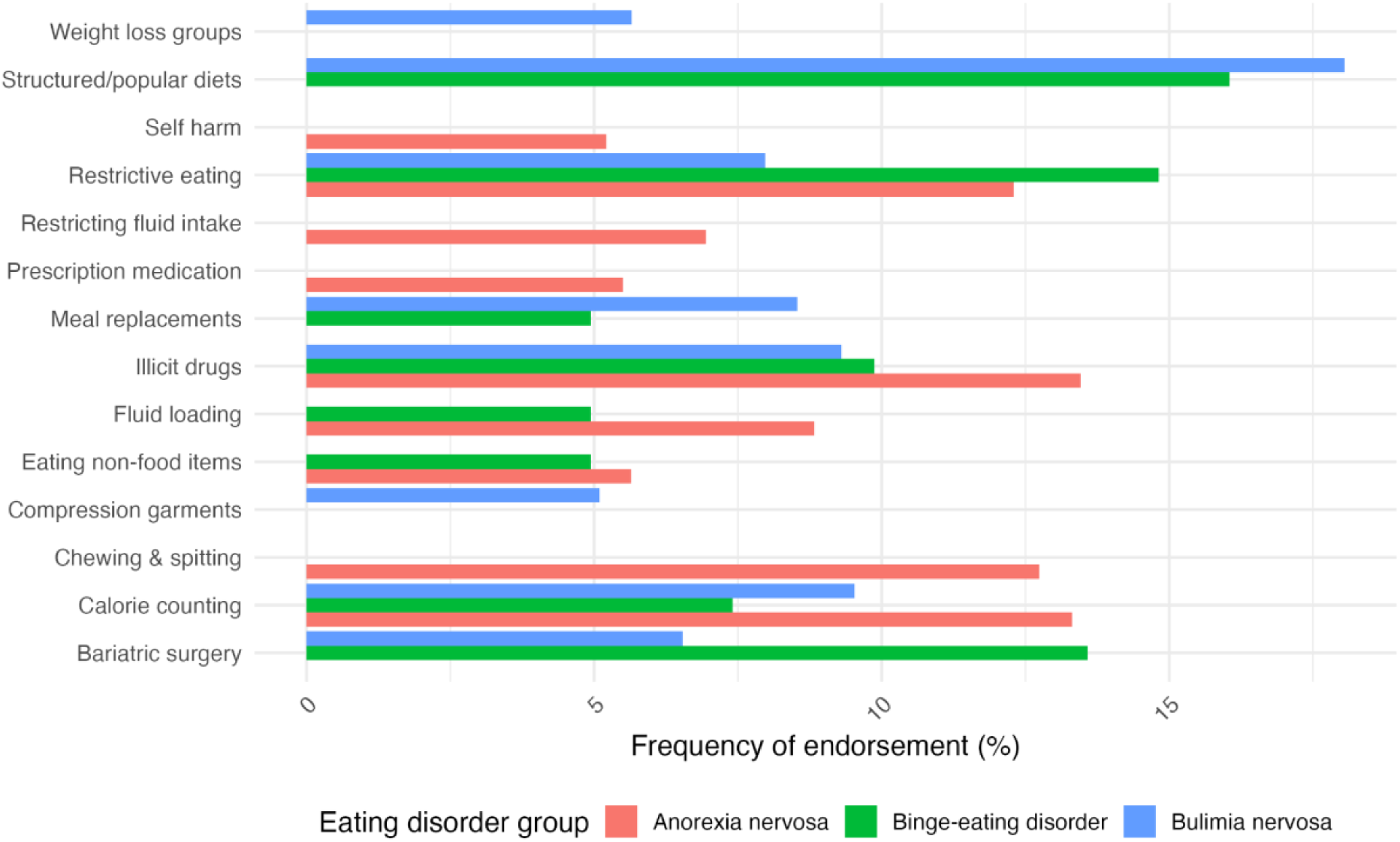
Frequencies of weight loss behaviours by eating disorder: anorexia nervosa (across subtypes; *n* = 691), bulimia nervosa (*n* = 903), and binge-eating disorder (*n* = 81). If the bar is not displayed, the weight loss behaviour was not endorsed.

We also focussed on men (*n* = 87), of whom 17 (19.5%) identified as transgender men. Of these self-identified men, 56 (64.4%) reported to be homosexual, bisexual, or asexual. After data cleaning, 99 tokens were analysed. Men endorsed weight loss behaviours most frequently closely mirroring the broader sample (**Table 2**).

## Discussion

In 2025, our study represents the largest systematic analysis of weight loss behaviours that may be overlooked by diagnostic interviews and questionnaires for eating disorders. We identified four themes–restriction-based approaches, medical interventions, body manipulation, and food avoidance–through text mining over 3,000 tokens derived from free-text responses. The open-ended format of the two questions in the ED100K.V3 and the questionnaire’s completion in private encouraged more candid disclosures from participants. This resulted in rich, authentic data reflecting behaviours people may hesitate to share in clinical settings.

Our findings highlight a crucial issue, that individuals with eating disorders often engage in weight loss behaviours that are not included in the current diagnostic criteria. Standard eating disorder assessment tools, such as the EDE-Q 6.0 and the EPSI, evaluate restriction-based behaviours like restrictive eating, calorie counting, and detox teas, and medical interventions like the use of steroids (Fairburn & Beglin, 1994; Forbush et al., 2013). The semi-structured interview EDE examines food restrictions (i.e., limitations on quantity, type, and specific food rules) as well as the use of diuretics, laxatives, self-induced vomiting, and excessive exercise. It also assesses other noteworthy dysfunctional weight-control behaviours (e.g., spitting, insulin underuse, thyroid medication misuse), requesting details on their frequency (number of days) and nature. However, these tools do not account for the full range of behaviours identified in our study, such as the use of non-food items, extreme food manipulation techniques, or off-label drug use for weight control. This gap in the diagnostic framework can lead to missed or incorrect diagnoses, as patients may not disclose certain behaviours unless specifically prompted, often due to the secrecy and shame surrounding eating disorders (Barko & Moorman, 2023). Without a comprehensive understanding of all relevant behaviours, clinicians may overlook critical aspects and health risks. For example, consuming non-food items may create digestive complications and off-label use of prescription medications poses considerable health risks like electrolyte disturbances or heart problems (Hendricks, 2017; Kariuki et al., 2016).

A key finding was that 81 participants with binge-eating disorder engaged in weight loss and compensatory behaviours typically linked to bulimia nervosa but not included in binge-eating disorder criteria. This challenges the clear-cut boundaries between eating disorders. The endorsement of restriction-based approaches by those with binge-eating disorder aligns with research as bariatric surgery may be pursued as a weight loss method according to national treatment guidelines for obesity (da Luz et al., 2018). Additionally, individuals with binge-eating disorder may adopt risky, overlooked weight loss methods, such as eating non-food items, meal replacements, and stimulant drugs, potentially worsening illness severity.

One of our study’s strengths lies in its hypothesis-free qualitative approach encouraging participants to candidly describe their behaviours, allowing for the identification of a broad range of weight loss behaviours not currently covered by diagnostic criteria. Additionally, our study is strengthened by the high number of participants. The study has the following limitations. A key consideration is the clarity of the free-text question asking participants to disclose any additional methods used to control their body shape or weight. For example, several responses described strategies to gain weight, such as marijuana to encourage appetite and protein shakes. Since both the GLAD Study and EDGI UK are ongoing, this question will be refined.

Research suggests that adolescents from minoritised ethnic groups demonstrate similar or higher tendencies to adopt weight loss behaviours compared with white adolescents (Goldschmidt et al., 2008). As our UK sample is mostly white, female, heterosexual, and highly educated, future research should recruit participants from more varied racial, ethnic, gender, and socioeconomic backgrounds. The GLAD Study and EDGI UK have already launched strategic advertising to enhance diversity. Future research should conduct in-depth interviews to understand motivations, context, and evolution of these weight loss behaviours.

## Conclusion

This study uncovered a wide range of weight loss behaviours that are often overlooked by current research and diagnostic frameworks, highlighting the limitations of existing tools. Developing psychometric tools designed to capture these broader behaviours could further enhance detection and facilitate earlier intervention. Future research should aim to diversify the sample and employ detailed interviews to gain a deeper understanding of these behaviours, improving the inclusivity of diagnostic criteria.

## Data Availability

Data from the GLAD Study and EDGI UK are not publicly available however are available via a data request application to the NIHR BioResource (https://bioresource.nihr.ac.uk/using-our-bioresource/academic-and-clinical-researchers/apply-for-bioresource-data/)

## Ethics statement

The London - Fulham Research Ethics Committee approved the GLAD Study on 21st August 2018 (REC reference: 18/LO/1218) and EDGI UK on 29th July 2019 (REC reference: 19/LO/1254) following a full review. The NIHR BioResource has been approved as a Research Tissue Bank by the East of England – Cambridge Central Committee (REC reference: 17/EE/0025).

## Acknowledgements

We gratefully acknowledge the participation of all National Institute for Health and Care Research (NIHR) BioResource, the NIHR BioResource Centre Maudsley, Biomedical Research Centre at South London and Maudsley NHS Foundation Trust and King’s College London volunteers and thank the BioResource staff for their help with volunteer recruitment. We thank the NIHR Biomedical Research Centre at South London and Maudsley NHS Foundation Trust and King’s College London for funding. We thank all participants who have kindly taken part in this study. This study represents independent research supported by the NIHR Biomedical Research Centre BioResource at South London and Maudsley NHS Foundation Trust and King’s College London. The views expressed are those of the author(s) and not necessarily those of the NHS, NIHR, Department of Health and Social Care or King’s College London.

## Author contributions

**Conceptualization:** Saakshi Kakar, Helena L. Davies, Christopher Hübel, Una Foye, Moritz Herle, Gerome Breen

**Data curation:** Saakshi Kakar, Helena L. Davies, Christopher Hübel, Gursharan Kalsi, Chelsea M. Malouf, Iona Smith, Laura Meldrum

**Formal analysis:** Saakshi Kakar, Helena L. Davies, Christopher Hübel, Elisavet Palaiologou

**Funding acquisition:** Gerome Breen, Gursharan Kalsi

**Investigation:** Saakshi Kakar, Helena L. Davies, Christopher Hübel, Gerome Breen, Una Foye, Moritz Herle

**Methodology:** Saakshi Kakar, Helena L. Davies, Christopher Hübel, Gerome Breen, Una Foye, Moritz Herle,

**Project administration:** Saakshi Kakar, Helena L. Davies, Christopher Hübel, Gursharan Kalsi, Gerome Breen, Chelsea M. Malouf, Iona Smith, Laura Meldrum

**Resources:** Saakshi Kakar, Gursharan Kalsi, Chelsea Mika Malouf, Saakshi Kakar, Gerome Breen, Chelsea M. Malouf, Iona Smith, Laura Meldrum

**Software**: Saakshi Kakar, Helena L. Davies, Christopher Hübel

**Supervision**: Christopher Hübel, Gerome Breen, Helena L. Davies, Una Foye, Moritz Herle

**Validation**: Saakshi Kakar, Christopher Hübel, Helena L. Davies, Elisavet Palaiologou, Chelsea M. Malouf, Laura Meldrum

**Visualisation**: Saakshi Kakar, Una Foye, Moritz Herle, Christopher Hübel

**Writing – original draft:** Saakshi Kakar, Una Foye, Moritz Herle

**Writing – review & editing:** Saakshi Kakar, Helena L. Davies, Chelsea M. Malouf, Gursharan Kalsi, Christopher Hübel, Gerome Breen, Una Foye, Moritz Herle, Elisavet Palaiologou, Laura Meldrum, Iona Smith, Karina L. Allen

